# A Digital CRISPR-based Method for the Rapid Detection and Absolute Quantification of Viral Nucleic Acids

**DOI:** 10.1101/2020.11.03.20223602

**Authors:** Xiaolin Wu, Cheryl Chan, Yie Hou Lee, Stacy L. Springs, Timothy K. Lu, Hanry Yu

**Author notes:** Correspondence should be addressed to Timothy K. Lu and Hanry Yu.

## Abstract

Quantitative real-time PCR and CRISPR-based methods detect SARS-CoV-2 in 1 hour but do not allow for the absolute quantification of virus particles, which could reduce inter-lab variability and accelerate research. The 4-hour reaction time of the existing digital PCR-based method for absolute virus quantification is too long for widespread application. We report a RApid DIgital Crispr Approach (RADICA) for the absolute quantification of SARS-CoV-2 DNA and Epstein–Barr virus DNA in human samples that yields results within 1 hour. For validation, we compared RADICA to digital PCR for quantifying synthetic SARS-CoV-2 DNA and Epstein–Barr viral DNA. RADICA allows absolute quantification of DNA with a dynamic range from 0.6 to 2027 copies/µL (R^2^ value > 0.98), without cross-reactivity on similar virus or human background DNA. Thus, RADICA can accurately detect and quantify nucleic acid in 1h without thermal cycling, providing a 4-fold faster alternative to digital PCR-based virus detection.

## Introduction

Methods to detect virus, as well as to quantify viral load, are needed for diagnostics, therapeutics, and vaccines to combat the worldwide spread of infectious disease, such as COVID-19. The reverse transcription-polymerase chain reaction (RT-qPCR) is considered a gold standard for viral infection diagnosis. However, quantification via RT-qPCR relies on the use of external standards or references, and the results can be variable, with a 20–30% variability reported even within trained laboratories^1-3^. Thus, an absolute quantification method with improved precision and accuracy is vital for virus research^4-6^.

Digital PCR is increasingly being used as a highly accurate and sensitive method for the absolute quantification of nucleic acids^1, 7, 8^. In a digital PCR reaction, the PCR mixture is separated into thousands of individual reactions, resulting in the amplification of either zero or one of the nucleic acid target molecules present in each partition. Since the PCR reaction in each partition proceeds independently, absolute quantification by digital PCR is more precise than RT-qPCR and more tolerant of inhibitors; furthermore, digital PCR overcomes poor amplification efficiency^1^. The sensitivity and precision of digital PCR-based viral detection have been demonstrated in quantitative detection and viral load analysis of SARS-CoV-2-infected patient samples with reduced inter-lab variability and fewer false negatives and fewer false positives compared with RT-PCR^6, 9, 10^. In addition to its application in viral diagnostics, digital PCR has also been applied to other areas of virus research, including the study of the aerodynamic transmission of SARS-CoV-2^5^. The main drawback of digital PCR, however, is the relatively long reaction time (∼4 hours) needed as a result of the 1-2°C/s ramp up/down rate for efficient inter-partition heat transfer during thermal cycling, compared to that of qPCR, which requires 1 hour. Reducing the reaction time of digital PCR is therefore crucial in enabling the adoption of the technology for rapid virus detection in hospitals and clinics^11^.

Isothermal amplification methods, which amplify the nucleic acid target molecule at a constant temperature and thereby reduce the reaction time, have also been used for viral detection. These include methods that employ recombinase polymerase amplification (RPA) or loop-mediated isothermal amplification (LAMP)^12, 13^. More recently, innovative diagnostic methods using RNA-guided CRISPR/Cas system have been developed to detect nucleic acids. In the RNA-guided CRISPR/Cas system, Cas effectors such as Cas12a and Cas13a are exploited for their collateral cleavage activity, which refers to the degradation of other nonspecific DNA/RNA oligos such as fluorescently-tagged reporter oligos, once the Cas protein finds and cleaves a specific DNA/RNA target^14, 15^. By combining RPA- or LAMP-mediated isothermal amplification of the target molecule with the CRISPR/Cas biosensing system, methods such as SHERLOCK and DETECTR have detected dengue virus and human papillomavirus, as well as SARS-CoV-2, in clinical samples^16-22^. However, as CRISPR-based methods are not quantitative and require multiple manipulations between the amplification and detection steps, there remains a need for a quantitative, rapid, and robust viral detection method.

Here, we report the development of a digital CRISPR method for the rapid, sensitive, and specific detection of viral nucleic acids at a constant temperature. This method combines the advantages of quantitative digital PCR, rapid isothermal amplification, and specific CRISPR detection into a one-pot reaction system that partitions the individual reactions into 10,000 compartments on a commercial high-density chip. In this study, we demonstrate an optimized RApid DIgital Crispr Approach (RADICA) that allows for absolute quantification of viral nucleic acids at a constant temperature in one hour. We validated this method using DNA containing the N (nucleoprotein) gene of SARS-CoV-2 and showed a linear signal-to-input response of R^2^ value > 0.99. We compared our RADICA detection system against the traditional digital PCR method and show that the RADICA system (1h vs 4h) was faster and had sensitivity and accuracy comparable to that of traditional digital PCR. Also, this method is highly specific and do not have cross activity on other similar virus or human background DNA. We also used RADICA in the absolute quantification of Epstein–Barr virus from human B cells (R^2^ value > 0.98). Our rapid and sensitive RADICA allows for the accurate detection and absolute quantification of viral nucleic acids in one hour.

## Results

### Design of RADICA

Commercial chips for sample partitioning and matched fluorescence reader for endpoint detection were used in RADICA^23^. In this system, each CRISPR-based reaction mix is sub-divided into 10,000 partitions on the chip, resulting in an average partition volume of 1.336 nL. We first optimized the bulk CRISPR reaction to achieve a one-copy-per-1.336 nL partition detection sensitivity on the chip. This is equivalent to femtomolar detection sensitivity in a bulk reaction. We selected the Cas12a homolog from *Lachnospiraceae bacterium* ND2006 (LbCas12a) as it showed the highest signal-to-noise ratio relative to other Cas12a homologs from a previous study^17^. To test if the RADICA could detect viral DNA with femtomolar sensitivity without pre-amplification, serially-diluted dsDNA (double stranded DNA) was incubated with LbCas12a together with its CRISPR RNA (crRNA) and a reporter (quenched fluorescent DNA). The sensitivity of detection using the CRISPR-based method without pre-amplification in a bulk reaction was found to be 10 pM (Supplementary Fig. 1), which did not meet the femtomolar sensitivity requirement of RADICA.

**Figure 1.**
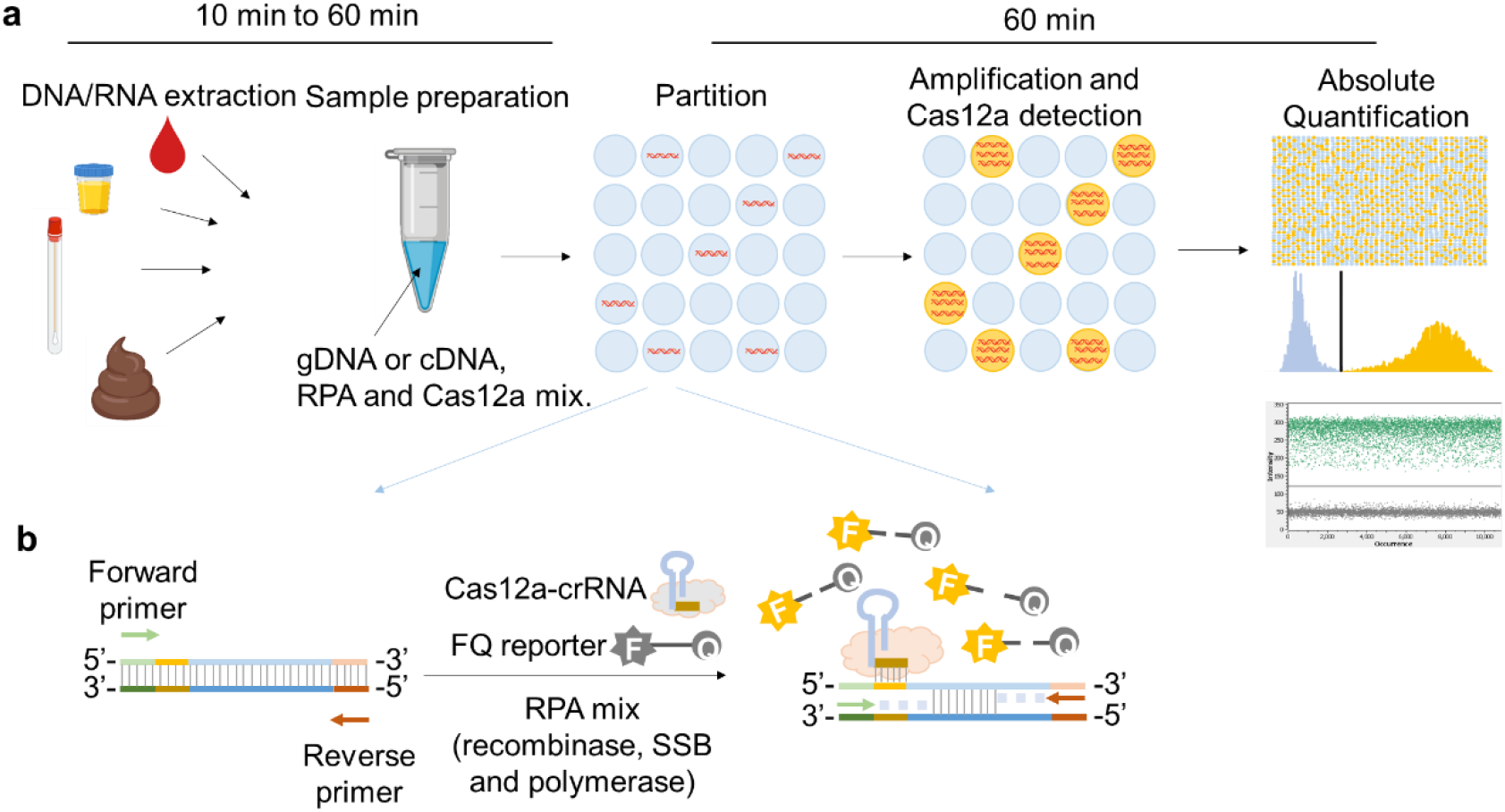
Schematic illustration of RADICA. **a**, The workflow of RADICA sample partitioning on a chip for absolute quantification of nucleic acid targets. Generally, after the DNA/RNA extraction step, different kind of clinical samples can be used for detection and quantification of various targets. The sample mixture containing DNA/cDNA, RPA reagents, and Cas12a-crRNA-FQ probes is distributed randomly into thousands of partitions. In each partition, the DNA is amplified by RPA and detected by Cas12a-crRNA, resulting in a fluorescent signal in the partition. Based on the proportion of positive partitions and on Poisson distribution, the absolute copy number of the nucleic acid target is quantified. **b**, Illustration of RPA-Cas12a reaction in each positive partition. In each partition containing the target nucleic acid, the primers bind to the target nucleic acid and initiate amplification with the aid of recombinase and DNA polymerase. Because of the strand displacement of DNA polymerase, the exposed crRNA-targeted ssDNA sites are bound by Cas12a-crRNA complexes. Cas12a is then activated and cleaves the nearby FQ reporters to produce a fluorescence readout.

To increase the sensitivity of detection of the CRISPR-based method, an isothermal amplification step was used. RPA was chosen for the isothermal amplification step because its reaction temperature (25°C to 42°C) is compatible with that of Cas12a (25°C to 48°C). This allowed for a one-step digital RPA-CRISPR absolute quantification method that eliminates multiple manipulations inherent in two-step CRISPR-based detection methods such as SHERLOCK and DETECTR^14, 15^. To avoid Cas12a-mediated cleavage of the target molecule before amplification, we designed the crRNA to target single-stranded DNA (ssDNA) that is generated only after amplification of the target molecule (Fig. 1b)^22^. Another advantage of this method is the ease of designing ssDNA-targeting crRNA over traditional dsDNA-targeting crRNA, because the nuclease activity of Cas12a on ssDNA has been reported to be independent of the presence of protospacer adjacent motif (PAM)^24^.

The RADICA developed in this study is illustrated in Fig. 1a. Extracted DNA samples are loaded onto on the chip by capillary action, and the reaction is partitioned into 10,000 compartments, resulting in zero or one target molecule in each compartment. To prevent spontaneous target amplification by RPA at room temperature^25^, the RPA-CRISPR reaction was prepared without the addition of Mg^2+^, which is required for the polymerase activity. All reactions were prepared on ice and samples were loaded within one minute to prevent premature target amplification. The partitioned reactions were incubated in isothermal water baths, heat blocks, or warm rooms.In each compartment containing the target molecule (Fig. 1b), RPA initiates from one DNA strand and subsequently exposes the crRNA-targeted ssDNA region on the other strand. As the amplification proceeds, Cas12a cleaves the positive ssDNA strand, triggering its collateral cleavage activity, which in turn cleaves the proximal quenched fluorescent probe (ssDNA-FQ reporter) to generate a fluorescence signal. At the same time, ongoing amplification of the other DNA strand exponentially amplifies the target DNA, triggering more Cas12a activation and increasing the fluorescence readout. The proportion of positive-to-negative compartments is analyzed based on the endpoint fluorescence measurement, and the copy number of the target nucleic acid is calculated based on the Poisson distribution, allowing for absolute quantification of the sample (Fig. 1a). Concurrent detection of compartments in each tube by the Clarity™ Reader shortens detection time to within minutes.

### RADICA optimization

To validate and optimize the RADICA, G-block DNA or plasmids containing the SARS-CoV-2 N (nucleoprotein) gene region were used and primers and crRNAs specific for the SARS-CoV-2 N gene were designed accordingly based on previous studies^22^. The target regions overlap with those of the China CDC assay (N gene region) with some modification to meet the primer and crRNA design (Supplementary Table 1). To optimize the Cas12a-mediated reaction, a bulk reaction using 0.1 nM and 1 nM dsDNA as a target was performed with a range of Cas12a/crRNA concentrations. We found that in the presence of a constant amount of target DNA and probe, comparable fluorescence signal intensities were detected between 50 nM to 250 nM Cas12a-crRNA concentrations, suggesting that changing the Cas12a/crRNA concentration did not influence the reaction (Supplementary Fig. 2).

**Figure 2.**
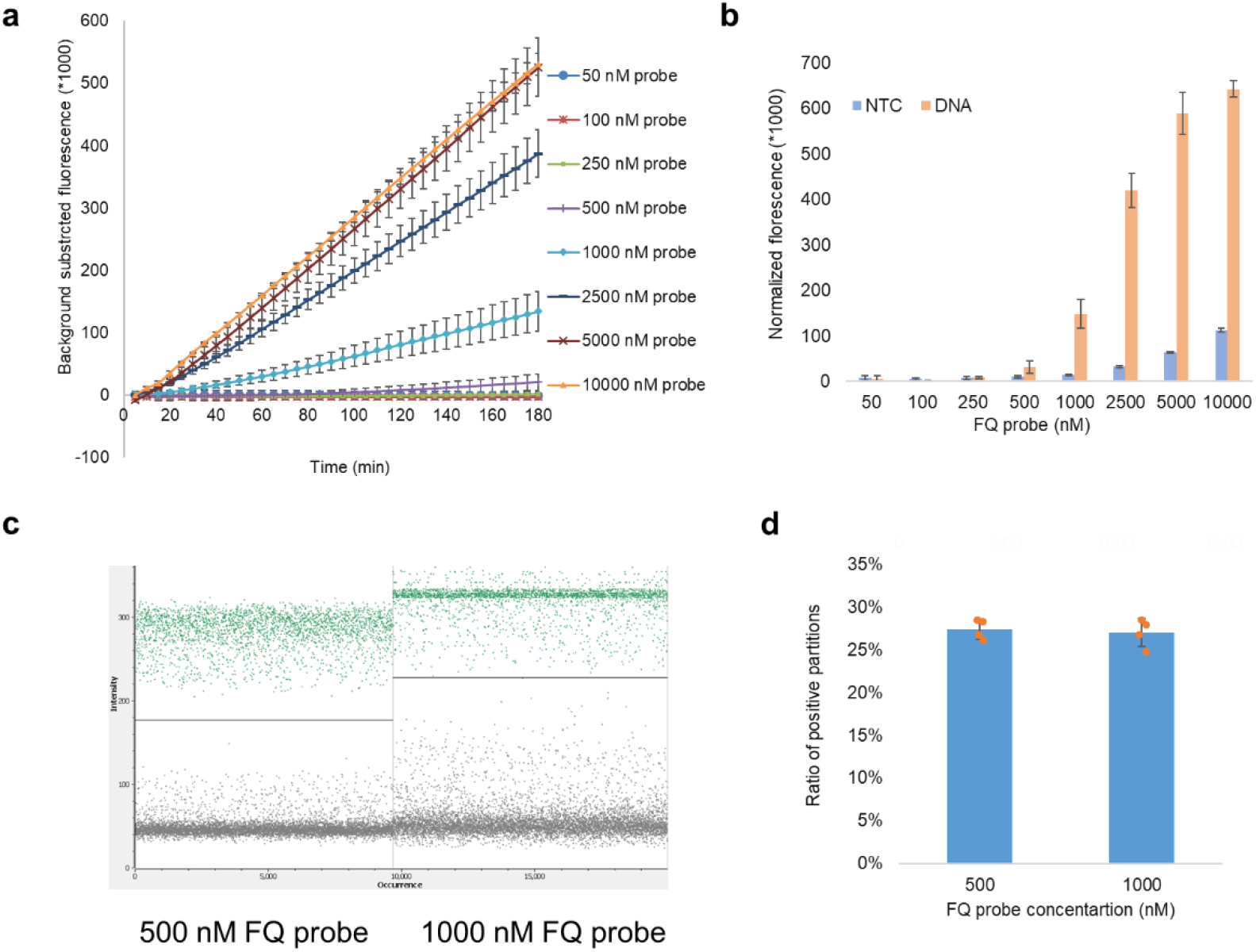
Optimization of FQ probe concentration in RADICA. **a, b**, Cas12a reaction in bulk reactions with different FQ probe concentrations in the presence or absence of a constant concentration (0.1 nM) of target DNA. **a**, Time course reaction of Cas12a with FQ probes at concentrations ranging from 50 nM to 10,000 nM. X-axis indicates the reaction time; y-axis indicates the background-subtracted fluorescence signal. **b**, Fluorescence signal of DNA and non-template control obtained with FQ probes at concentrations ranging from 50 nM to 10,000 nM. **c, d**, RADICA reaction with the same concentrations of target DNA but different probe concentrations. **c**, Fluorescence intensity of the negative partitions (background noise) and positive partitions (positive signals) on the chip obtained with FQ probes at concentrations of 500 or 1000 nM. **d**, Histogram showing ratios of positive partitions on the chip with FQ probes, at concentrations of 500 or 1000 nM, in the presence of target DNA (4 replicates for each FQ probe concentration).

Since the quenched fluorescent probe is another key component that influences the reaction, we optimized the FQ assay by incubating increasing amounts of FQ probes with constant concentrations of Cas12a-crRNA and target DNA. As expected, CRISPR-mediated fluorescence signal intensities increased with increasing amounts of FQ probes (from 250 nM to 5 µM), although higher probe concentrations also resulted in higher background noise (Fig. 2a,b). At FQ probe concentrations above 5 µM, the signal-to-noise ratio could not be further enhanced (Fig. 2b). To ensure that the fluorescence signal generated on the partitioned chip was within the reader’s detection range, different FQ probe concentrations were tested in independent digital CRISPR reactions in the presence of the target DNA and the fluorescence measured on the digital PCR fluorescence reader. We found that in the presence of the same target DNA, the proportions of positive partitions were comparable regardless of the FQ probe concentration used (Fig. 2d).

However, only the background noise and positive signals generated in the reaction with 500 nM FQ probe concentration were within the reader’s detection range, while the reactions containing 1000 nM FQ probe concentration yielded higher background noises, which are difficult to separate from positive signals (Fig. 2c). We therefore used 500 nM FQ probe concentrations to achieve high signal-to-noise ratios for subsequent experiments.

An additional optimization step involved developing a one-pot reaction that combines the RPA and Cas12a reactions. We performed the bulk reaction at 25°C, 37°C and 42°C, which are temperatures within the reaction temperature ranges of RPA (25°C to 42°C) and Cas12a (25°C to 48°C). First, we tested the reaction using serial dilutions of plasmid DNA at 25°C and 42°C. The reaction proceeded at both of these reaction temperatures, with a limit of detection of about 9.4 copies/µL. However, at 25°C, the reaction proceeded significantly more slowly with lower positive signals and a higher background than the reaction performed at a 42°C (Supplementary Fig. 3). Next, we assessed the effect of different temperatures (25°C, 37°C and 42°C) on reactions containing a constant amount of plasmid DNA (37.5 copies/µL). We found that higher temperatures accelerated the reaction (Supplementary Fig. 3c). Taken together, our results suggest that 42°C is the optimal temperature for the RPA-Cas12a reaction.

**Figure 3.**
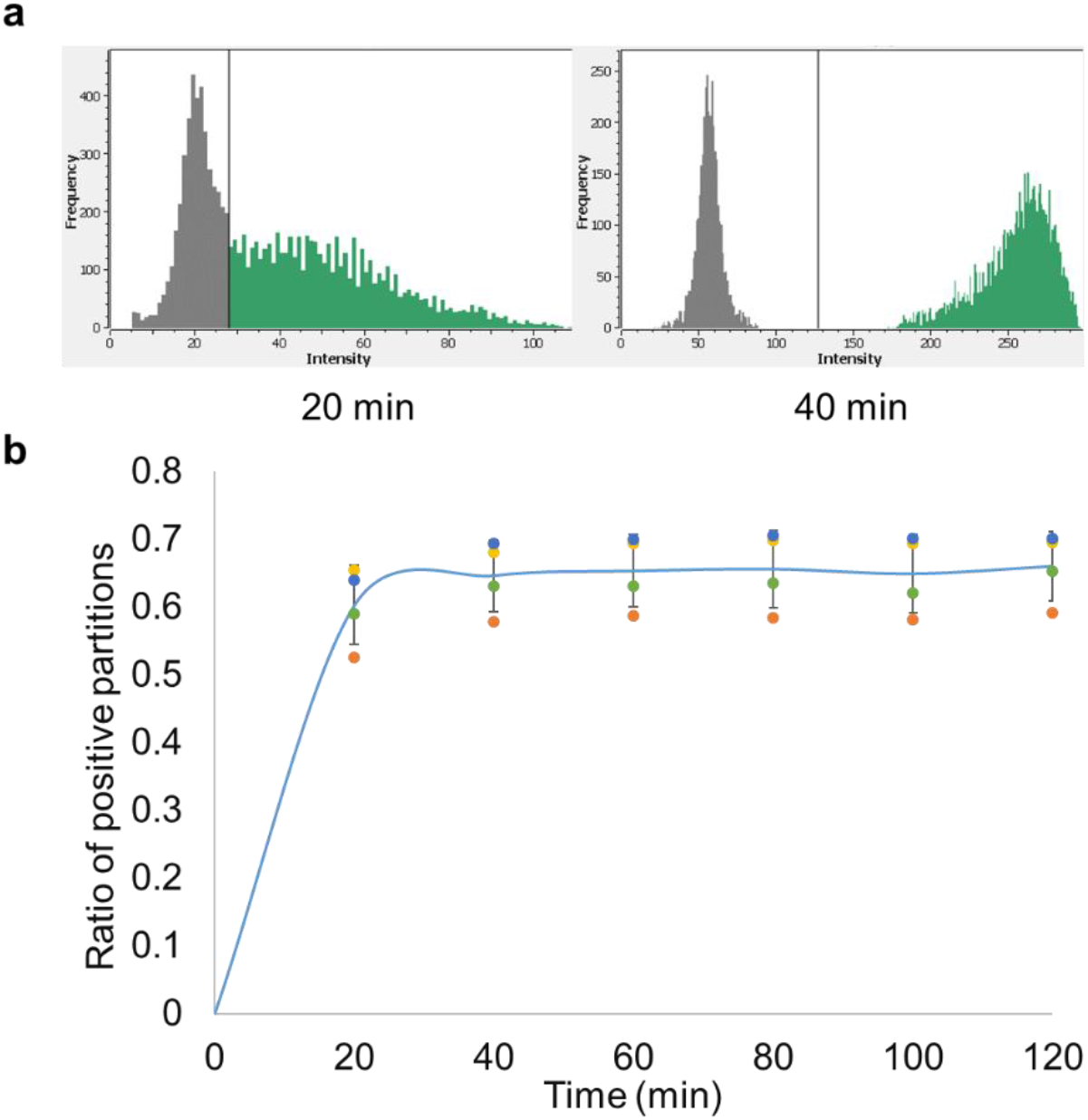
Time course reaction of RADICA. **a**, Fluorescence intensity of the partitions on the chip at two time points. The x-axis represents fluorescence intensity while the y-axis represents the frequency of the partitions. The left peak (low fluorescence level; dark grey) on the fluorescence intensity histogram represents the negative partitions while the right peak (high fluorescence level; green) indicates the positive partitions. As the CRISPR reaction proceeds, the fluorescence levels of the positive partitions increase and the right peak shifts further to the right. **b**, The proportion of positive partitions at different time points of RADICA. Each DNA replicate is represented by a data point with a unique color. Starting at about 60 minutes, the fluorescence signal plateaus and the ratio of positive partitions reaches a stable level.

We next investigated whether the reaction time affected the precision of RADICA on plasmid DNA detection at 42°C. As shown in Fig. 3a, the reaction proceeded quickly with some fluorescence signal detected in several compartments at 20 min, but with a low signal-to-noise ratio at this time point. As the reaction proceeded, two distinct peaks indicating the negative (left) and positive (right) partitions were detected at 40 min (Fig. 3a). Analysis of the ratio of positive partitions on the chip at the different time points revealed that the number of positive partitions reached a plateau after 60 min in all four replicates, suggesting that 60 min was the earliest end-point measurement (Fig. 3b). All subsequent experiments were therefore performed for 60 minutes.

### Absolute quantification of SARS-CoV-2 DNA using RADICA

We next characterized the assay performance of RADICA in detecting and quantifying SARS-CoV-2 and compared it to that of digital PCR. In this assay, linearized plasmid containing the SARS-CoV-2 N gene was serially diluted and used as the target DNA in the aforementioned optimized RADICA or digital PCR reactions. Using RADICA-based detection, a proportional increase in the number of positive partitions was observed with increasing concentrations of the target DNA (Fig. 4a), indicating the good quantitative performance of RADICA. Although few partitions in the negative control were classified as having a positive signal due to non-specific amplification, the average of 10 negative control replicates give us an average of 0.165 copies/µL readout and the limit of blank (LoB) is 0.413 copies/µL, which is about half of our limit of detection (LoD), 0.897 copies/µL (Fig. 4a, Supplementary Table 2).

**Figure 4.**
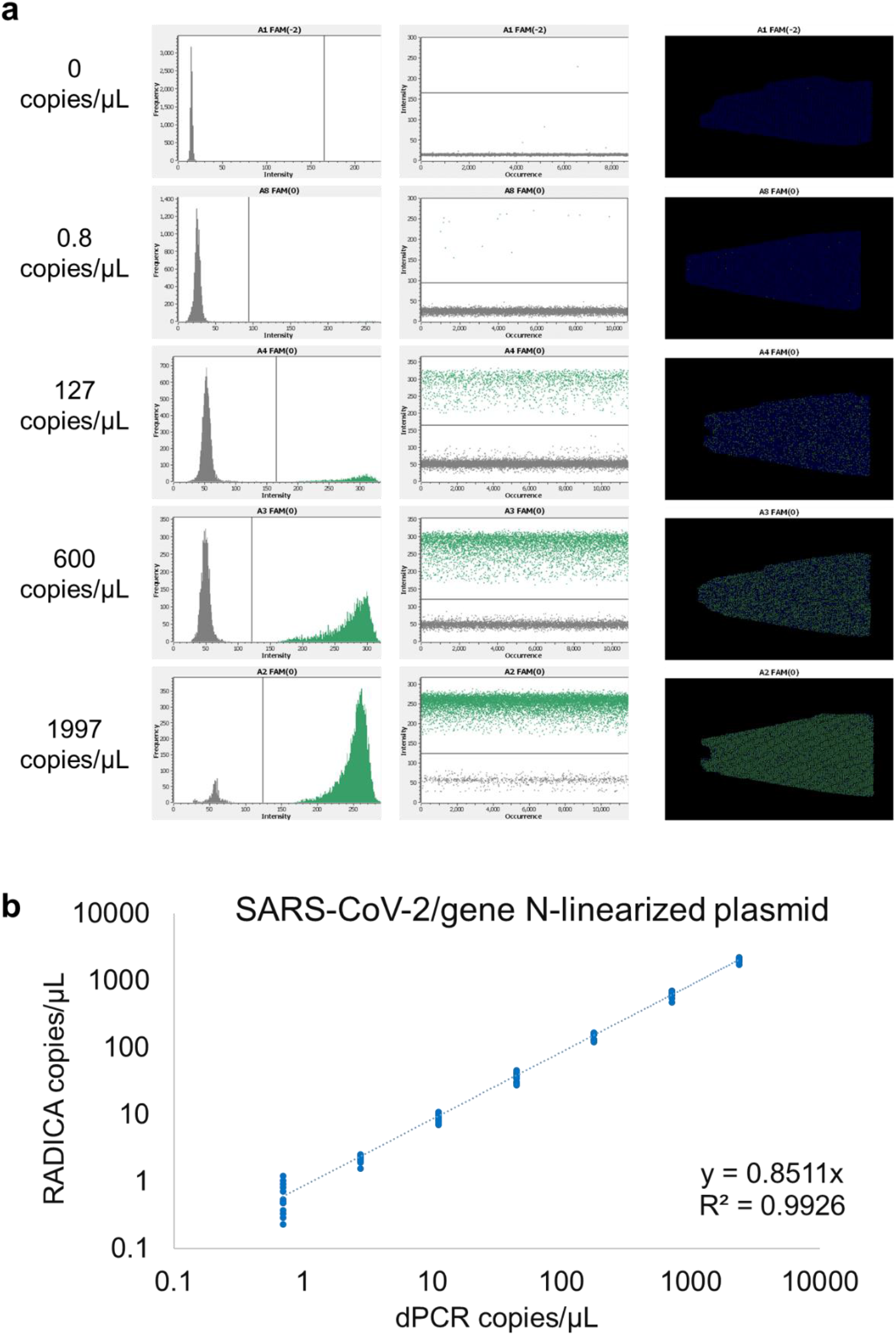
RADICA-based detection of different concentrations of SARS-CoV-2 N gene DNA. **a**, Fluorescence intensity histogram, scatter plot, and position plot of the partitions on the chip for serial dilutions of DNA. Four dilutions of linearized plasmid DNA encoding the SARS-CoV-2 N gene (0.8, 127, 600, 1997 copies/µL) and one non-template control (without plasmid DNA) were used as input DNA. The x-axis represents fluorescence intensity while the y-axis represents the frequency of the partitions. The left peak (low fluorescence level; dark grey) on the fluorescence intensity histogram represents the negative partitions while the right peak (high fluorescence level; green) indicates the positive partitions. In the scatter plot and position plot, each dot represents one partition on the chip. Green dots represent positive partitions with a high fluorescence level while grey or blue dots correspond to negative partitions with a low fluorescence level. **b**, Comparison of the absolute quantification result of RADICA and digital PCR. Each point represents one sample. The original linearized plasmid DNA concentration was measured by using Clarity™ digital PCR and diluted to different concentrations (x-axis). The diluted DNA was then measured by using the RADICA. The calculated RADICA DNA concentrations are plotted on the y-axis.

To test the robustness and reproducibility of RADICA, at least ten independent RADICA reactions using the SARS-CoV-2 N gene as the target DNA were performed on different days. The coefficient of variation (CV) observed for most samples was ≤15% except for the lowest dilution (0.6 copies/µL), indicating the limit of quantification (LoQ) of this method is around 2.2 copies/µL of the viral genome (Supplementary Table 2). To assess the accuracy of RADICA-based nucleic acid detection against that of digital PCR, DNA concentrations measured by RADICA were plotted against the corresponding DNA concentrations obtained by digital PCR. Linear regression analysis revealed an R^2^ value of above 0.99 across a dynamic range from 0.6 to 2027 copies/µL, suggesting that RADICA was reliable for the absolute quantification of nucleic acids (Fig. 4b). These data highlight the sensitivity, accuracy, and speed of the digital CRISPR-based detection method developed in this study for the absolute quantification of nucleic acids in samples.

### Accuracy analysis of RADICA-based quantification on circular plasmid

Plasmids are routinely used as reference DNA or standards in many analytical DNA measurements. However, conformational changes in supercoiled DNA have been reported to have a profound effect on PCR-based quantification^26-28^. Unsuccessful single-molecule amplification of non-linearized plasmids was reported in a PCR-based study, which resulted in an underestimation for circular plasmid quantification on some dPCR machines^29, 30^. To test whether plasmid conformation also affects the accuracy of RADICA, undigested plasmid containing the SARS-CoV-2 N gene was serially diluted and used for digital PCR or RADICA reactions. Concentrations of non-linearized plasmids measured by digital PCR were about half of those detected for linearized plasmids (regression coefficients at 0.5261) (Fig. 5d), which is in accordance with previous studies and indicates that the accuracy of digital PCR is influenced by plasmid conformation. Compared to digital PCR, RADICA showed higher amplification efficiency of the supercoiled plasmid DNA, as evidenced by the higher positive compartments ratio detected (Fig. 5a,b).

**Figure 5.**
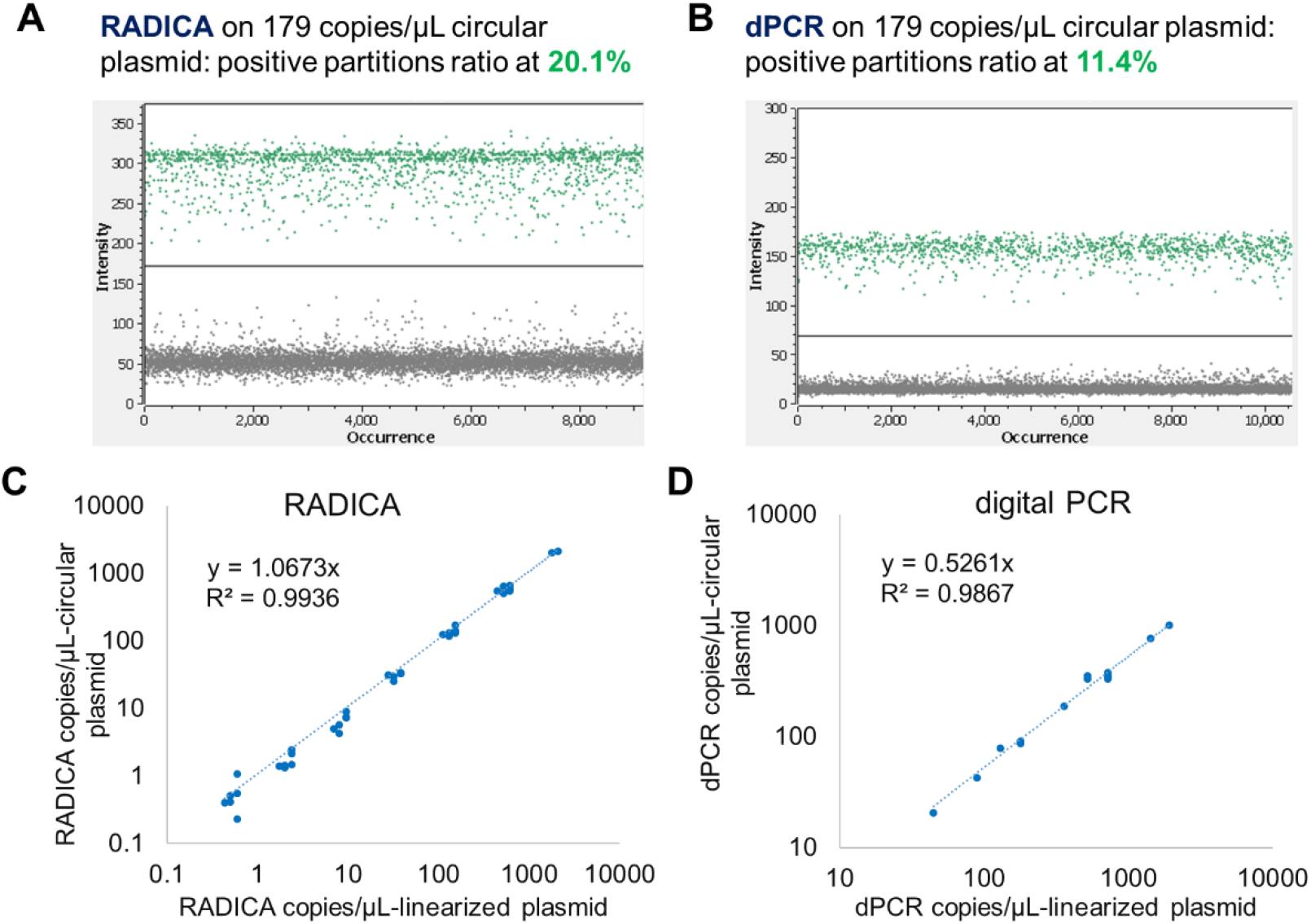
The effect of plasmid conformation on the accuracy of RADICA and digital PCR (dPCR). **a, b**, The positive and negative partitions of RADICA (**a**) and dPCR (**b**) on detection of 179 copies/µL circular plasmid. **c, d**, Comparison of the absolute quantification result for linearized plasmid and circular plasmid of RADICA (**c**) and dPCR (**d**).

RADICA concentrations of non-linearized plasmids were highly concordant with those of linearized plasmids (regression coefficients at 1.0673) (Fig. 5c), suggesting that the accuracy of RADICA is not affected by plasmid conformation.

### Specificity analysis of RADICA-based detection

Primer and crRNA designs are key in determining the specificity of CRISPR-based nucleic acid detection assays. Previous studies have shown the ability of RPA to tolerate up to nine nucleotide base-pair mismatches across primer and probe binding sites^25^. To specifically detect SARS-CoV-2 using RADICA, primers and crRNAs must be designed to specifically bind the SARS-CoV-2 target DNA and not its closely-related coronaviruses, such as MERS-CoV and other related human coronaviruses. We first analysed the binding sites of the primers and crRNAs that were originally designed based on the consensus sequence of the genome of 264 SARS-CoV-2 strains, available on the GISAID database^22, 31, 32^. The consensus sequence of these SARS-CoV-2 target regions was aligned with corresponding regions of SARS-CoV-2-related beta coronaviruses, such as SARS-CoV, MERS-CoV, and human coronaviruses Human-CoV 229E/HKU1/NL63/OC43. No cross-binding regions were observed with the other SARS-CoV-2-related coronavirus analyzed (Supplementary Fig. 4a). A comparison between the binding site sequences of SARS-CoV-2 and its most similar relative, SARS-CoV, showed that there were 13 sequence variations across the primer and crRNA binding sites (three, two, and seven sequence variations at the forward primer, reverse primer, and crRNA binding site, respectively). This is more than the nine nucleotide base-pair mismatch tolerance threshold for RPA, which therefore predicts the specificity of the designed primers and crRNA for SARS-CoV-2-specific CRISPR-based detection. To test the specificity of the reaction, we assayed the bulk RPA-Cas12a reaction using target plasmids containing the complete N gene from SARS-CoV-2, SARS-CoV, and MERS-CoV (Supplementary Fig. 4b,c). Positive fluorescence signals were observed only in the reaction containing the SARS-CoV-2 plasmid, not in reactions containing SARS-CoV and MERS-CoV plasmids (Supplementary Fig. 4b,c). The absence of cross-reactivity with the other related coronaviruses tested in this study validates the specificity of the CRISPR assay for SARS-CoV-2.

### Background human DNA tolerance analysis of RADICA

Previous studies have reported that RPA reactions could be inhibited by high concentrations of background human DNA^33, 34^. We therefore first tested the RPA-Cas12a bulk reaction in the presence of various concentrations of background human DNA (Supplementary Fig. 5). In an RPA-Cas12a reaction with 37.5 copies/µL of target DNA, background human DNA concentrations below 2 ng/µL did not affect the reaction (Supplementary Fig. 5a).

Concentrations of background human DNA above 5 ng/µL in a bulk RPA-Cas12a reaction showed reduced fluorescent signal intensities, which is in agreement with the inhibitory concentrations of background DNA reported in previous studies using bulk RPA reactions^34^ (Supplementary Fig. 5a).

We also tested for possible inhibitory effects of background DNA on reactions carried out in small partitions. In an RPA-Cas12a reaction with 400 copies/µL of target DNA, 1 ng/µL of background human DNA (equivalent to about 4350 human cells per reaction) did not affect the RADICA reaction (Supplementary Fig. 5b). We also observed inhibition of the reaction containing 2 ng/µL of background human DNA, and complete inhibition of the reaction containing >5 ng/µL of background human DNA (Supplementary Fig. 5b). Nevertheless, since input DNA concentrations used for RADICA-based detection are typically below 1 ng/µL, our findings suggest that background DNA will not inhibit the RADICA reaction of samples within the dynamic range to be used for testing.

Previous studies have also reported that the tolerance of RPA for background DNA is dependent on target DNA concentrations present in the reaction^33, 34^. We therefore tested the effect of 1 ng/µL of background human DNA (equivalent to about 4350 human cells per reaction) on RADICA reactions with various concentrations of target DNA (Supplementary Fig. 5c). Our results show that 1 ng/µL of background DNA did not affect reactions that contained target DNA concentrations within the dynamic range of digital PCR detection, i.e., 0.6 to 2027 copies/µL (Supplementary Fig. 5c). Our findings confirm that the presence of background human DNA in the sample is not likely to affect the absolute quantification of RADICA.

### RADICA detection and absolute quantification of SARS-CoV-2 RNA

As SARS-CoV-2 is an RNA virus, we next tested whether RADICA could be combined with reverse transcription (RT) in a one-pot reaction for the absolute quantification of RNA. RNA corresponding to the SARS-CoV-2 gene N target region was synthesized using a T7 promoter-tagged PCR product and T7 RNA polymerase, and different concentrations of RNA were tested in bulk RT-RPA-Cas12a reactions. Our results show a lower-than-expected sensitivity of the one-pot RT-RPA-Cas12a bulk reaction, with a detection limit at 244 copies/µL (Supplementary Fig. 6b). To assess if this decrease in sensitivity was in part due to an inefficient reverse transcription process in the one-pot reaction, we employed two reverse primers to facilitate the reverse transcription reaction and increase the sensitivity of the one-pot reaction. We detected an increase in sensitivity of 61 copies/µL when two reverse primers were used (Supplementary Fig. 6c).

**Figure 6.**
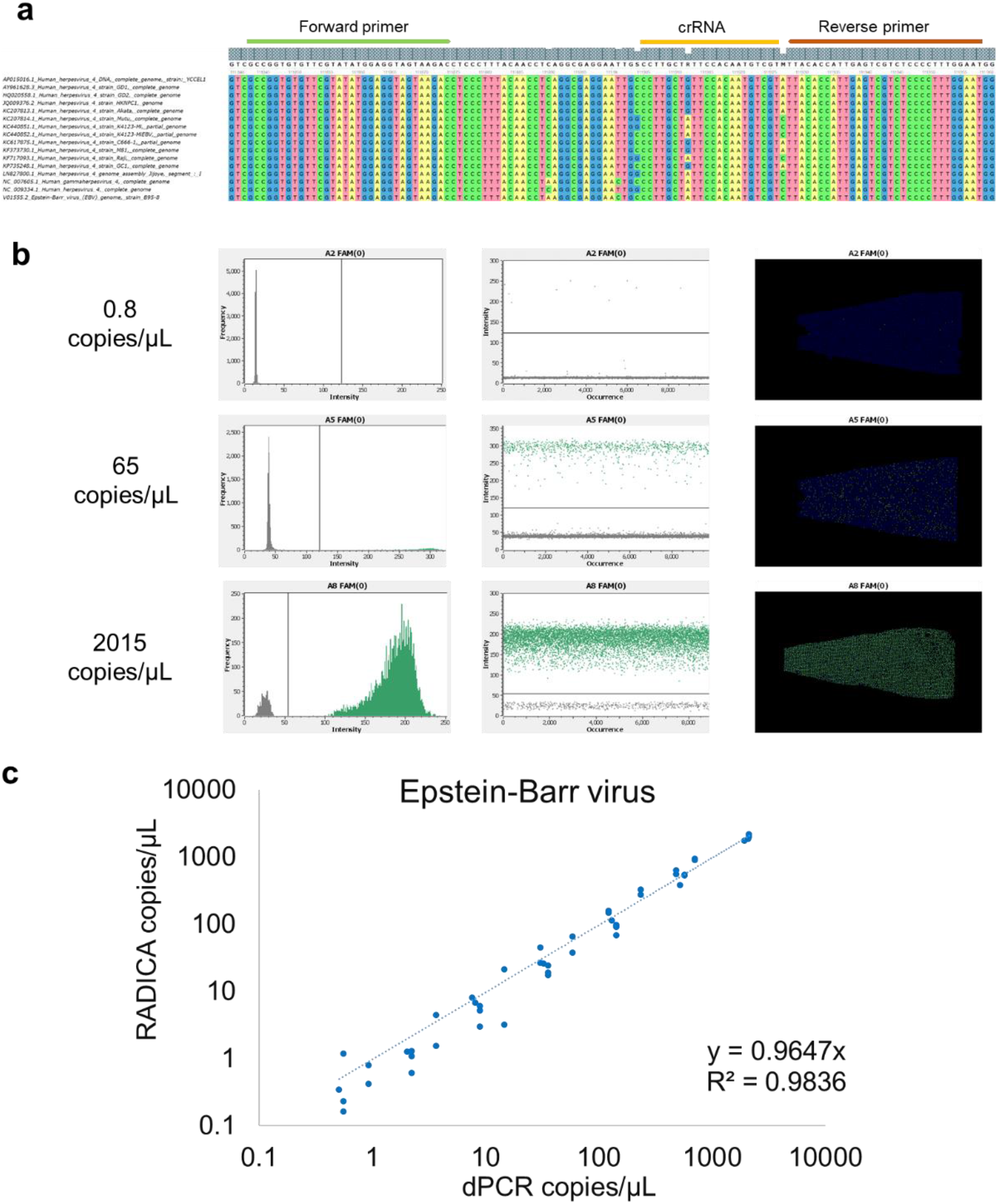
Absolute quantification of Epstein–Barr virus (EBV) by RADICA. **a**, Primer and crRNA design for RADICA assay specific for EBV. **b**, Fluorescence intensity histogram, scatter plot, and position plot of the partitions on the chip on serially-diluted EBV DNA. **c**, A comparison of the absolute quantification values obtained from RADICA and digital PCR methods using various concentrations of EBV DNA.

We tested the sensitivity of the one-pot RT-RPA-Cas12a reaction in small partitions on RADICA using varying concentrations of RNA. Notably, although the positive partition proportion increased with an increase in the concentration of input RNA, 1 copy of input RNA resulted in an increase of only 0.0177 copy as calculated by RADICA, suggesting that the RADICA does not accurately quantify RNA (Supplementary Fig. 6d). Although the addition of two reverse primers increased the signal, it was insufficient to achieve a one-copy-per-partition sensitivity (Supplementary Fig. 6e). Our findings suggest that absolute quantification of RNA by RADICA may require prior conversion to cDNA before the sample is analysed on a digital chip.

### Absolute quantification of Epstein–Barr virus via RADICA

Having demonstrated the accuracy of RADICA on SARS-CoV-2 DNA samples, we tested the ability of our RADICA method to perform absolute quantification of Epstein–Barr virus (EBV) DNA samples. To design primers and crRNA that were universal to both type I and type II EBV, the genomes of 16 EBV strains were analysed to identify conserved regions across all 16 EBV strains. A conserved DNA region within the Epstein-Barr nuclear antigen 1 (EBNA1) was used as the target sequence (Fig. 6a). Viral DNA extracted from chemically-induced EBV-harboring human B cells was diluted to concentrations ranging from 0.5 to 2100 copies/µL, and used as the target DNA in both RADICA and digital PCR reactions. In the RADICA-based detection, samples loaded in the partition chip were incubated for 1 h at 42 °C, followed by endpoint fluorescence detection and copy number determination. Notably, the positive partition signal increased with an increase in the concentration of input EBV DNA (Fig. 6b). The copy numbers measured by RADICA were in full agreement (R^2^ value > 0.98) with those measured by digital PCR (Fig. 6c). Our findings validate the accuracy and sensitivity of our RADICA for the absolute quantification of viral DNA rapidly within an hour, which is a 4-fold reduction in reaction time compared to digital PCR-based detection.

## Discussion

In our study, we have developed a rapid and accurate digital CRISPR method for the absolute quantification of viral DNA. The performance characteristics of this method were validated using SARS-CoV-2 synthetic DNA and EBV DNA, and compared to those of absolute quantification digital PCR method, the current gold standard. Our RADICA achieved sensitivity and detection limits (LoD 0.897 copy/µL) comparable with those of qPCR and other isothermal methods, such as SHERLOCK and DETECTR, with the ability of absolute quantification (Supplementary Table 3). The significant advantage of RADICA over dPCR is its speed: RADICA can perform absolute quantification rapidly within an hour, which is four times faster than current dPCR-based detection.

Absolute quantification for viral detection could not only facilitate clinical processing but also benefit research. Viral loads closely parallel transmission risk and disease severity. High SARS-CoV-2 viral loads have been reported to correlate with the course of infection and mortality^6, 35-38^. These reports underscore the urgent need for rapid and sensitive virus detection and quantification methods to monitor viral load as the basis for clinical decision making. Such methods are also needed for mechanistic studies, transmission studies, vaccine development, and therapeutics for COVID-19. Although there currently exist many diagnostic methods available for virus detection, these methods usually do not allow for a rapid and precise quantification of the viral load (Supplementary Table 3).

RADICA reported in the study is four times faster than the traditional digital PCR-based methods used for the absolute quantification of nucleic acids. Additionally, the isothermal feature of RADICA-based detection assay confers faster amplification of the viral target using a simple constant-temperature heat bath, enabling rapid viral detection that can be deployed even in low-resource areas. In recent years, other digital isothermal methods, such as RPA- or LAMP-based digital PCR methods, have been developed for detecting a variety of DNA targets^39-41^. However, these methods are limited by their low specificity, due to the inherent tolerance of RPA/LAMP-based methods for base-pair mismatches as compared to traditional PCR methods^42-44^. Our RADICA overcomes this by exploiting the specificity conferred by the Cas12a-crRNA-based targeting system. The collateral cleavage activity of Cas12a amplifies the signal and thus increases the sensitivity.

Another advantage of RADICA over other CRISPR-based methods^16-21^ is its one-pot reaction design, which reduces manual manipulation and increases reproducibility. In this streamlined one-pot reaction, both nucleic acid amplification and CRISPR-based detection are combined into a single step in a closed tube, significantly reducing the risk of cross-contamination between samples during batch processing. A major drawback of current CRISPR-based methods is the complexity of designing appropriate crRNAs that are limited to target regions in proximity to a PAM. This limitation may potentially complicate CRISPR-based virus detection since mutations in the viral PAM sequence may disable recognition by the Cas protein as the virus evolves. In contrast, our simpler digital CRISPR crRNA design is independent of the PAM sequence because it targets single-stranded DNA generated after amplification^24^.

RADICA reported here uses commercially available chips and devices that can potentially be adapted to other devices already in use at some hospitals and service laboratories. These are QuantStudio 3D Digital PCR System (Thermo Fisher), QIAcuity Digital PCR System (QIAGEN), and Droplet Digital PCR System (Bio-Rad). We therefore envisage greater ease of adoption of our technology at these facilities. Furthermore, RADICA offers a potentially customizable solution that is amenable to other DNA isothermal amplification platforms such as loop-mediated isothermal amplification, rolling circle amplification, and strand displacement amplification technologies, as well as the use of other Cas proteins, such as Cas13a, Cas12b, Cas14 for multiplex detection.

We have established and characterized RADICA, which combines the speed and sensitivity of isothermal amplification, the specificity of CRISPR-based detection, and the ability to obtain absolute quantification by sample partitioning. Our RADICA detects a concentration of viral DNA as low as 0.897 copy/µL and enables rapid, absolute quantification with a dynamic range of 0.6 to 2027 copies/µL within one hour at a constant temperature, with no cross-reactivity to other similar viruses. Future work will focus on expanding the applications of RADICA to areas such as gene expression analysis, rare mutant detection, copy number variation, and sequencing library quantification. Applications of such rapid analytics will also benefit cell therapy, pharmaceutical, environmental, public health, security and food industry to potentially determine the replication competency of adventitious agents.

## Materials and methods

### Materials

#### Preparation of primers and DNA targets

Oligonucleotides (primers), ssDNA-FQ reporters, SARS-CoV-2 N gene-containing G-Block, SARS-CoV-2, SARS-CoV, and MERS N gene-containing plasmids were synthesized by or purchased from Integrated DNA Technologies. The SARS-CoV-2 N gene-containing plasmid was linearized using FastDigest ScaI (Thermo Scientific) and then used as DNA target. The SARS-CoV-2 N gene-containing plasmid was used as a template to amplify the N gene using primer N-RNA-F/ N-RNA-R by Platinum™ SuperFi II PCR Master Mix (Invitrogen). The PCR product was purified by QIAquick PCR Purification Kit (QIAGEN) and used as RNA synthesis template.

#### Synthetic RNA target preparation

Since N-RNA-F has a T7 promoter sequence, the amplified DNA using N-RNA-F/R primer will contain a T7 promoter upstream of gene N. The T7 tagged N gene dsDNA was transcribed into SARS-CoV-2 RNA using HiScribe™ T7 High Yield RNA Synthesis Kit (New England Biolabs) according to the manufacturer’s protocol. The synthesized RNA was purified using Monarch® RNA Cleanup Kit (New England Biolabs) after treatment with DNase I (RNase-free, New England Biolabs).

#### crRNA preparation

Constructs were ordered as DNA from Integrated ssDNA Technologies with an appended T7 promoter sequence. crRNA ssDNA was annealed to a short T7 primer (T7-3G IVT primer^45^ or T7-Cas12scaffold-F^46^) and treated with fill-in PCR (Platinum™ SuperFi II PCR Master Mix) to generate the DNA templates. These DNA were used as templates to synthesize crRNA using the HiScribe™ T7 High Yield RNA Synthesis Kit (New England Biolabs) according to published protocols^45, 46^. The synthesized crRNA was purified using Monarch® RNA Cleanup Kit (New England Biolabs) after treatment with DNase I (RNase-free, New England Biolabs), Thermolabile Exonuclease I (New England Biolabs), and T5 Exonuclease (New England Biolabs).

### Primer and crRNA design

SARS-CoV-2 primers and crRNA were designed based on previously published papers^22^ or 264 SARS-CoV-2 genome sequences from GISAID^31,32^. Other human-related coronavirus sequences were downloaded from NCBI. UGENE software was used to analyze and align viral genomes (MUSCLE or Kalign). Consensus sequences (Threshold: 90%) of 264 SARS-CoV-2 genomes, 328 SARS-CoV, 572 MERS-CoV, 70 Human-CoV-229E genomes, 48 Human-CoV-HKU1 genomes, 71 Human-CoV-NL63, and 178 Human-CoV-OC43 were exported separately from UGENE and used for specificity analysis.

Epstein–Barr virus primers and crRNA were designed based on consensus sequences of 16 virus genomes including both type I and type II EBV (NCBI: AP015016.1, AY961628.3, HQ020558.1, JQ009376.2, KC207813.1, KC207814.1, KC440851.1, KC440852.1, KC617875.1, KF373730.1, KF717093.1, KP735248.1, LN827800.1, NC_007605.1, NC_009334.1, V01555.2).

### Digital PCR quantification

SARS-CoV-2 N gene quantification: The G-block, plasmid, dsDNA and RNA concentrations were quantified by digital PCR. Serial dilutions of targets were mixed together with 500 nM CHNCDC-geneN-F, 500 nM CHNCDC-geneN-R, 250 nM CHNCDC-geneN-P, 1x TaqMan™ Fast Virus 1-Step Master Mix (for RNA, Applied Biosystems) or TaqMan™ Fast Advanced Master Mix (for DNA, Applied Biosystems), 1x Clarity™ JN solution (JN Medsys). For RNA samples, the reaction mixture was incubated at 55°C 10 min before partitioning the reaction mix on Clarity™ autoloader. Then the reaction partitions were sealed with the Clarity™ Sealing Enhancer and 230 μL Clarity™ Sealing Fluid, followed by thermal cycling using the following parameters: 95 °C for 15 min (one cycle), 95 °C 50 s and 56 °C 90 s (40 cycles, ramp rate = 1 °C/s), 70 °C 5 min. The endpoint fluorescence of the partitions was detected using Clarity™ Reader and the final DNA copy numbers were analyzed by Clarity™ software.

EBV quantification: Serial dilutions of EBV DNA was used for dPCR quantification by Clarity™ Epstein-Barr Virus Quantification Kit (JN Medsys) according to the manufacturer’s protocol.

### Cas12a bulk assay without preamplification

Unless otherwise indicated, 50 nM EnGen® Lba Cas12a (New England Biolabs), 50 nM crRNA, and 250 nM FQ ssDNA probe were incubated with dsDNA dilution series in NEB buffer 2.1 at 37°C, and fluorescence signals were measured every 5 min.

### RPA-Cas12a bulk assay

The one-pot reaction combining RPA-DNA amplification and Cas12a detection was performed as follows: 300 nM forward primer, 300 nM reverse primer, 500 nM FQ probe, 1x RPA rehydration buffer containing 1 x RPA Pellet (TwistDx), 200 nM EnGen® Lba Cas12a (New England Biolabs), 200 nM crRNA, were prepared followed by adding various amounts of DNA input, and 14 mM magnesium acetate. When RNA was used as a target, 300 nM reverse primer 2 was used with 10 U/µL PhotoScript Reverse transcriptase (New England Biolabs) or 10 U/µL SuperScript™ IV Reverse Transcriptase (Invitrogen) and 0.5 U/µL RNase H (Invitrogen or New England Biolabs), as indicated. The reaction mixture was incubated at 42°C unless otherwise indicated and fluorescence kinetics were monitored every 1 min.

### RADICA quantification

The RADICA reaction was prepared by adding 1x Clarity™ JN solution (JN Medsys) to the RPA-Cas12a bulk reactions stated above. 15 µL of the mixture was loaded on the chip by a Clarity™ autoloader for sample partitioning. The reaction partitions were sealed with the Clarity™ Sealing Enhancer and 230 μL Clarity™ Sealing Fluid, followed by incubation at 42°C for 1 hour, unless otherwise indicated. After incubation, a Clarity™ Reader was used to read the fluorescent signal in the partitions, and Clarity™ software was used to calculate input DNA copy numbers.

### Limit of Blank (LoB), Limit of Detection (LoD), and Limit of Quantitation (LoQ) calculation

LoB, LoD, and LoQ were calculated based on the following equation^47^ using the statistics of RADICA quantification on linearized plasmid in 10 replications (Supplementary Table 2):

LoB = mean ^blank^ + 1.645 (SD ^blank^)

LoD = LoB + 1.645 (SD ^low concentration sample^)

LoQ = the lowest concentration of CV ≤ 20%

### Growing EBV-2 from Jijoye cells

Jijoye cells were treated with 4 mM sodium butyrate and 24 ng/ml tetradecanoyl phorbol acetate (TPA). Supernatants were harvested 4-5 days post-treatment by centrifugation at 4,000g for 20 min and passing over a 0.45 µm filter to remove cellular debris. Viral particles were pelleted by ultracentrifugation at 20,000 rpm for 90 min and resuspended in 1/100 the initial volume using complete RPMI or PBS if viruses were to be further purified. Concentrated viruses were further purified using OptiPrep gradient density ultracentrifugation at 20,000 rpm for 120 min, and the virus interface band collected and stored at -80°C for downstream analysis.

### Epstein–Barr virus DNA extraction

EBV DNA was extracted using QIAamp DNA Mini Kit (QIAGEN) according to the manufacturer’s protocol.

## Data Availability

More data are included in Supplementary Materials.

## Acknowledgments

We thank Karen Pepper (MIT) and Scott A. Rice(NTU) for careful editing and helpful comments on the manuscript. This research is supported by the National Research Foundation, Prime Minister’s Office, Singapore under its Campus for Research Excellence and Technological Enterprise (CREATE) programme, through Singapore MIT Alliance for Research and Technology (SMART): Critical Analytics for Manufacturing Personalised-Medicine (CAMP) Inter-Disciplinary Research Group.

## Author Contributions

X.W., T.K.L., and H.Y. designed the research. X.W. developed the RADICA, performed the experiment, and analyzed the data. C.C performed the Epstein–Barr virus culture and DNA extraction. Y.H.L., S.S., T.K.L., and H.Y. provided mentorship and feedback. X.W. wrote the original draft and all authors reviewed and edited the manuscript.

