## Supplementary information for "A Digital CRISPR-based Method for the Rapid Detection and Absolute Quantification of Viral Nucleic Acids"

[1]Singapore-MIT Alliance for Research and Technology, Critical Analytics for Manufacturing of Personalized Medicine Interdisciplinary Research Group, Singapore 138602, Singapore

[2]Center for Biomedical Innovation, Massachusetts Institute of Technology, Cambridge, MA, USA.

[3]Synthetic Biology Center, Massachusetts Institute of Technology (MIT), Cambridge, MA 02139, USA

[4]Synthetic Biology Group, Research Laboratory of Electronics, Massachusetts Institute of Technology (MIT), Cambridge, MA 02139, USA

[5]Broad Institute of MIT and Harvard, Cambridge, MA 02142, USA

[6]Department of Electrical Engineering and Computer Science, Massachusetts Institute of Technology (MIT), Cambridge, MA 02142, USA

[7]Harvard-MIT Division of Health Sciences and Technology, Cambridge, MA 02139, USA

[8]Department of Biological Engineering, Massachusetts Institute of Technology (MIT), Cambridge, MA 02142, USA

[9]Institute of Bioengineering and Nanotechnology, A*STAR, The Nanos, #04-01, 31 Biopolis Way, Singapore 138669, Singapore

[10]Mechanobiology Institute, National University of Singapore, T-Lab, #05-01, 5A Engineering Drive 1, Singapore 117411, Singapore

[11]Department of Physiology, The Institute for Digital Medicine (WisDM), Yong Loo Lin School of Medicine, MD9-04-11, 2 Medical Drive, Singapore 117593, Singapore

**Items included in supplementary information:**

**Supplementary Figures 1 to 6**

**Supplementary Tables 1 to 3**

**Supplementary References**

**Supplementary Table 1. Primers and probes used in this study.**

| Name | Sequence | Application |
|---|---|---|
| ssDNA-FQ reporter | /56-FAM/TTATT/3IABkFQ/ | Cas12a FQ reporter |
| N-RNA-F | gaaatTAATACGACTCACTATAgggATGTCTGATAATGGACCCCAAAAT | Amplify N gene on plasmid |
| N-RNA-R | gaaatTTAGGCCTGAGTTGAGTCAGCACT | Amplify N gene on plasmid |
| CHNCDC-geneN-F | GGGGAACTTCTCCTGCTAGAAT | dPCR Primer for N gene |
| CHNCDC-geneN-R | CAGACATTTTGCTCTCAAGCTG | dPCR Primer for N gene |
| CHNCDC-geneN-P | /56-FAM/TT GCT GCT G/ZEN/C TTG ACA GAT T/3IABkFQ/ | dPCR Probe for N gene |
| T7-3G IVT primer | GAAATTAATACGACTCACTATAGGG | univeral primer used for crRNA synthesis |
| N-N-RPA-F | AACTCCAGGCAGCAGTAGGGGAACTT | CRISPR Primer for N gene |
| N-N-RPA-R | CCTTTACCAGACATTTTGCTCTCAAG | CRISPR Primer for N gene |
| N-N-crRNA-IVT | AATCTGTCAAGCAGCAGATCTACACTTAGTAGAAATTACCCTATAGTGAGTCGTATTAATTTC | crRNA template for N gene, pair with T7-3G IVT primer |
| N-AIOD-F[1] | AGGCAGCAGTAGGGGAACTTCTCCTGCTAGAAT | RADICA Primer for N gene |
| N-AIOD-R[1] | TTGGCCTTTACCAGACATTTTGCTCTCAAGCTG | RADICA Primer for N gene |
| N-AIOD-crRNA-IVT-F | ggctggcaatggcggtgatgATCTACACTTAGTAGAAATTACCCTATAGTGAGTCGTATTAATTTC | crRNA template for N gene, pair with T7-3G IVT primer |
| T7-Cas12scaffold-F | gaaatTAATACGACTCACTATAGGTAATTTCTACTAAGTGTAGAT | univeral primer used for crRNA synthesis |
| N-RPA-RR | AGCAGATTTCTTAGTGACAGTTTGGCCTTGTTG | CRISPR Primer for N gene |
| N2-RPA-RR | AGCAAAATGACTTGATCTTTGAAATTTGGATCT | CRISPR Primer for N gene |
| N-JQ217-F[2] | CAACTTCCTCAAGGAACAACATTGCCAAAA | CRISPR Primer for N gene |
| N-JQ235-R[2] | TGGAGTTGAATTTCTTGAACTGTTGCGACT | CRISPR Primer for N gene |
| NF-CoV-RR | GAGAAGTTCCCCTACTGCTG | CRISPR Primer for N gene |
| NF-crRNA-1F | CTTCTACGCAGAAGGGAGCAatctacacttagtagaaatta | crRNA template for N gene, pair with T7-Cas12scaffold-F |
| EBV-EBNA1-F2 | GCCGGTGTGTTCGTATATGGAGGTAGTAAGAC | RADICA Primer for EBV |
| EBV-EBNA1-R2 | ATTCCAAAGGGGAGACGACTCAATGGTGTAA | RADICA Primer for EBV |
| EBNA-2R1-crRNAR | ACGACATTGTGGAAYAGCAAGGatctacacttagtagaaatta | crRNA template for EBV, pair with T7-Cas12scaffold-F |

**Supplementary Table 2. Quantification of linearized plasmid target by RADICA**

| Number of replications | dPCR measured concentration (copies/μL) | Mean of RADICA measured concentration (copies/μL) | Standard deviation (copies/μL) | Relative standard deviation or coefficient of variation (CV) |
|---|---|---|---|---|
| 10 | Non template control | 0.17 | 0.15 | - |
| 14 | 0.70 | 0.60 | 0.29 | 49.41% |
| 14 | 2.80 | 2.20 | 0.25 | 11.25% |
| 13 | 11.19 | 9.06 | 1.23 | 13.53% |
| 14 | 44.76 | 35.53 | 5.30 | 14.91% |
| 10 | 179.06 | 146.90 | 18.02 | 12.27% |
| 10 | 716.24 | 626.58 | 73.96 | 11.80% |
| 10 | 2387.46 | 2027.27 | 152.76 | 7.54% |

**Supplementary Table 3. Comparison of RADICA with other viral detection methods.**

| | RT-PCR | CRISPR-based isothermal method | RT-digital PCR | Digital RPA/digital LAMP | RADICA |
|---|---|---|---|---|---|
| **Detecting Substance** | RNA, DNA, cDNA | RNA, DNA, cDNA | RNA, DNA, cDNA | DNA, cDNA | DNA, cDNA |
| **LoD (for SARS-CoV-2)** | 5 copies per reaction | 20 copies per reaction | 2 copies per reaction | NA | 13.5 copies per reaction<br>(0.897 copies/µL in 15 µL reaction) |
| **Result Time** | 2 hours | **1 hour** | 4 hours | **1 hour** | **1 hour** |
| **Thermal cycling Requirements** | Yes | **No** | Yes | **No** | **No** |
| **All devices and regents commercially available** | **Yes** | **Yes** | **Yes** | No | **Yes** |
| **Ease of primer and crRNA design** | **Yes** | Moderate | **Yes** | Moderate | **Yes** |
| **Specificity** | **Good** | **Good** | **Good** | Normal | **Good** |
| **Absolute quantification** | No | No | **Yes** | **Yes** | **Yes** |

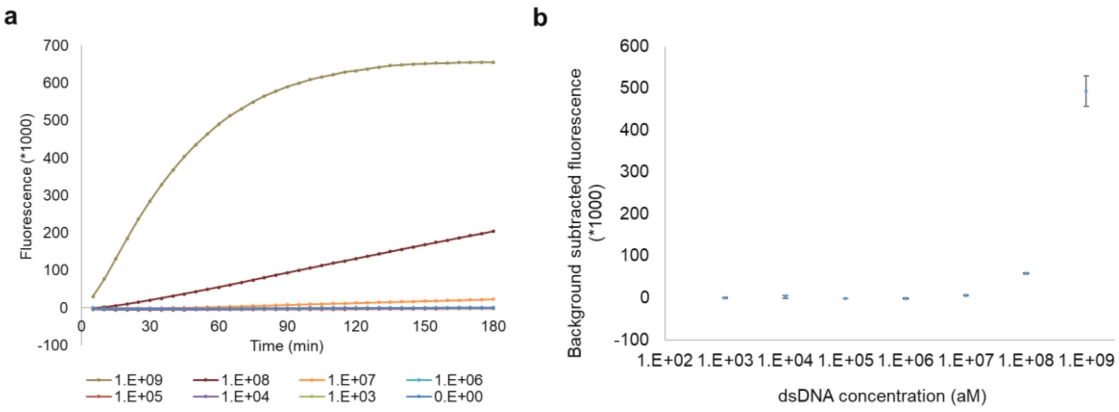


**Supplementary Figure 1. Cas12a detection sensitivity of dsDNA dilution series without pre-amplification. a,** Kinetics of Cas12a-crRNA on dsDNA dilution series. **b,** Detection sensitivity analysis of dsDNA dilution series with Cas12a-crRNA in 1h. 50nM Cas12-crRNA and 250 nM FQ-ssDNA probe were incubated with dsDNA dilution series at 37℃ and fluorescence was monitored every 5 min.

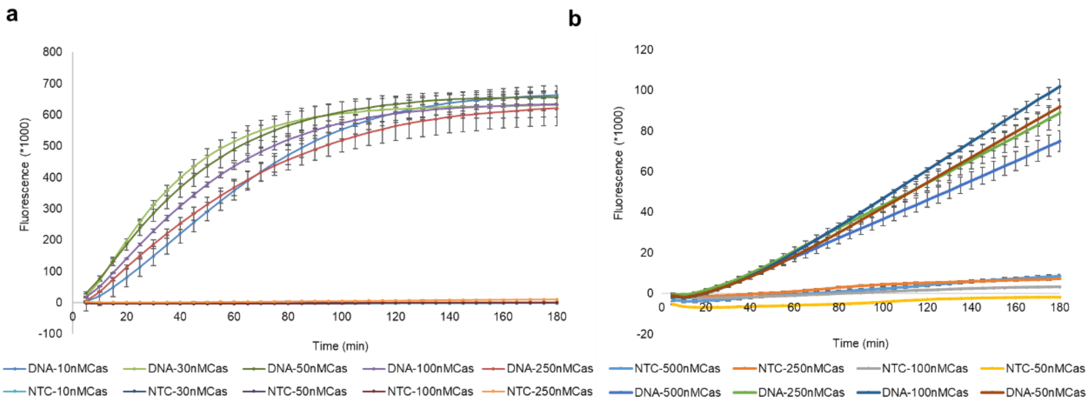


**Supplementary Figure 2. Detection of dsDNA without preamplification at different Cas12a/crRNA concentrations. a,** 1 nM dsDNA incubated with 10-250 nM Cas12a-crRNA and 250 nM FQ ssDNA probe. **b,** 0.1 nM dsDNA incubated with 50-500 nM Cas12a-crRNA and 500 nM FQ ssDNA probe.

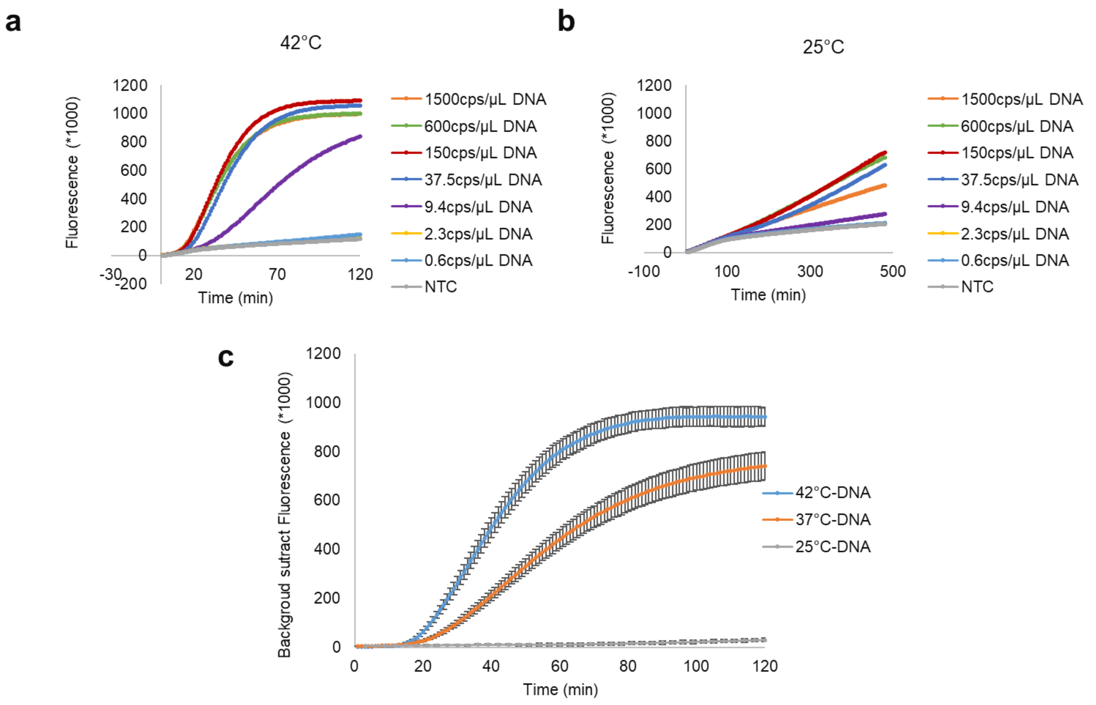


**Supplementary Figure 3. RPA+Cas12a bulk reaction at different temperatures. a, b,** RPA+Cas12a one-pot reaction on serial dilutions of DNA at 42°C (**a**) or 25°C (**b**). **c,** RPA+Cas12a one-pot reaction on 37.5 copies/μL plasmid DNA at different temperatures (25°C, 37°C, and 42°C).

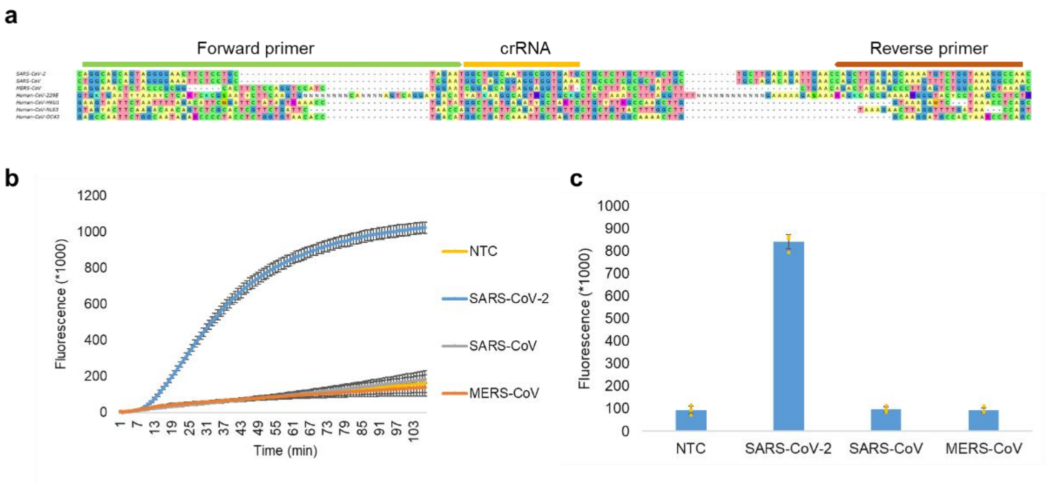


**Supplementary Figure 4. Specificity analysis for SARS-CoV-2. a,** Sequence alignment of the SARS-CoV-2 target region (N gene) and the corresponding regions on other human coronaviruses. **b,** Time course reaction of bulk RPA-Cas12a assay on SARS-CoV-2, SARS-CoV, and MERS-CoV N gene DNA target. The same concentration (25000 copies/μL) of the N gene target from different coronaviruses was tested by the bulk RPA-Cas12a assay. **c,** Specificity of the bulk RPA-Cas12a assay for detection of the SARS-CoV-2 N gene.

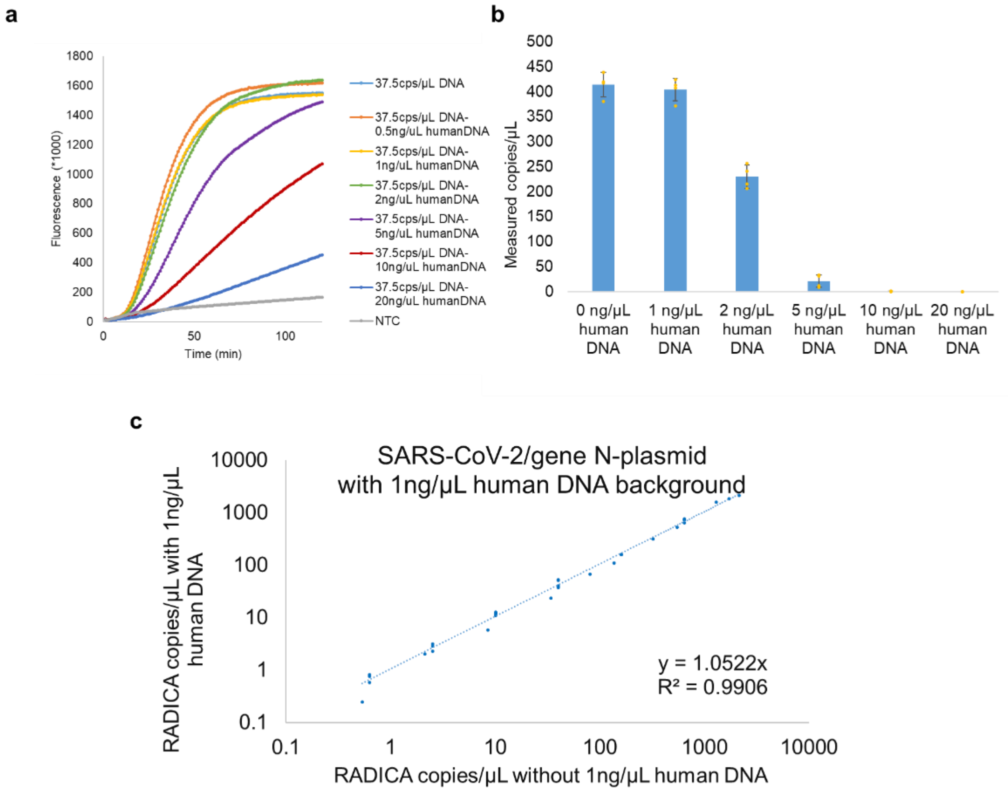


**Supplementary Figure 5. RPA inhibition by human background DNA. a,** Results of bulk RPA-Cas12a reaction to detect target DNA (i.e., the N gene from SARS-CoV-2) in the presence of various amounts of human background DNA. **b,** Results of RADICA reaction with target DNA and various amounts of human background DNA. Each point represents one sample. **c,** Comparison of the RADICA reaction with or without 1ng/μL human background DNA.

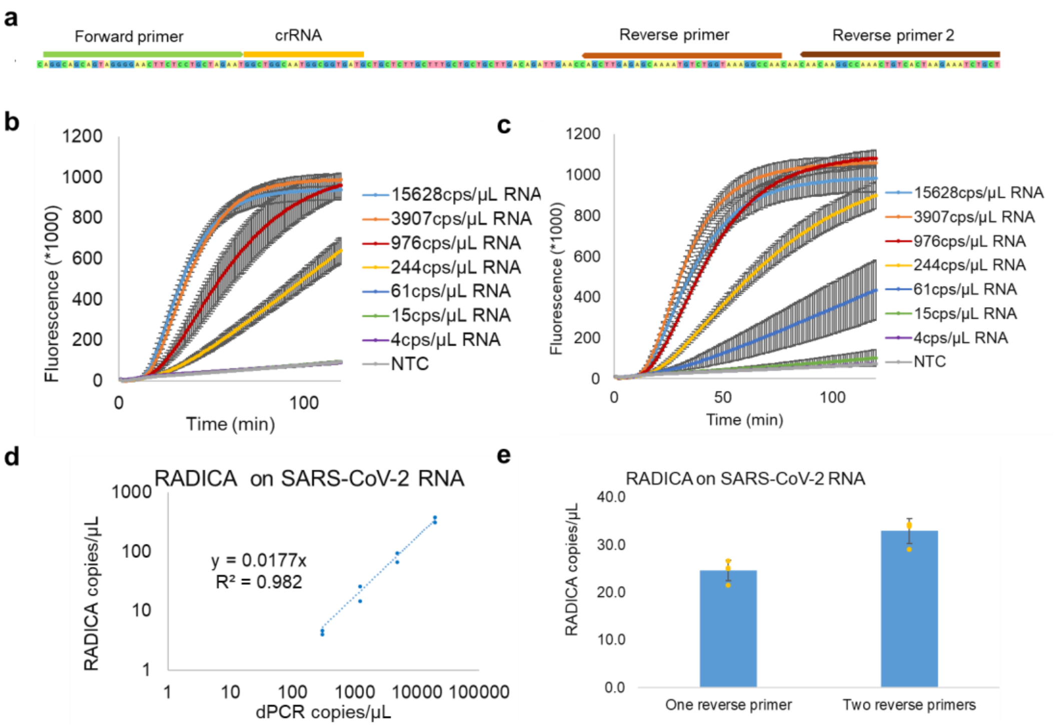


**Supplementary Figure 6. Digital isothermal CRISPR reaction on RNA samples. a,** Design of two reverse primers to increase the sensitivity. **b, c,** Bulk RPA-Cas12a reaction on RNA at different concentrations with normal one reverse primer design (**b**) or two reverse primers design (**c**). **d,** Comparison of the absolute quantification result of RADICA and digital PCR on SARS-CoV-2 RNA. **e,** Comparison of RADICA's performance on normal one reverse primer design and two reverse primers design.
